# Circulating Proteins as Predictors of Improvement in Physical Function in Hospitalized Older Adults with Geriatric Syndromes: Findings from the REHAB-HF Trial

**DOI:** 10.64898/2026.09.22.26363692

**Authors:** Abdulla A. Damluji, Scott A. Bruce, Christopher R. deFilippi

## Abstract

**Introduction:** Circulating proteins may augment prognostic scores and provide mechanistic insights for cardiovascular disease. Their role for adding precision to predicting response to therapeutic exercise training is less certain. Older adults with HF frequently experience geriatric syndromes, which may influence response to a rehabilitation intervention (RI). We sought to determine if the functional benefits of a 12-week tailored, structured, and progressive multidomain physical RI program in older adults, following a HF admission, could be estimated by pre-intervention levels of proteins.

**Methods:** Hospitalized adults age ≥ 60 years with HF were randomized to 12-week multidimensional (aerobic, strength, balance, flexibility) physical therapist-led RI versus attention control (AC). Outcomes included change in the short physical performance battery (SPPB) and 6-minute walk distance (6MWD). At pre-intervention and 12-weeks, 92 circulating proteins associated with CVD and inflammation were measured. Using linear regression, LASSO and ensemble tree-based approaches for flexible feature selection, proteins were selected for stratification of treatment response. Decision trees were developed to incorporate a multi-protein approach to optimize stratification of treatment response (effect measure modifiers [EMM]) for both functional outcomes. Differences in 12-week temporal changes based on RI vs AC and mediation analysis were used to assess possible causal roles for these proteins.

**Results:** Baseline proteomic data was available in 243/349 participants (69.6%). For SPPB, 2 proteins (GP6, ST2), and for 6MWD, 3 proteins (ALCAM, Gal-4, PECAM-1) were selected as EMMs. For SPPB, a decision tree with ST2 followed by GP6 level identified a differential 12-week improvement in score for RI vs AC from −0.8 (95%CI −1.9, 0.2) to 2.1 (95%CI 1.5, 2.7). For 6MWD, the optimized decision tree was based only on ALCAM with a differential 12-week improvement in score for RI vs AC from 10m (95%CI −17m, 36m) to 85m (95%CI 56m, 113m). However, none of the EMM proteins were mediators of change for the functional outcomes.

**Conclusions:** Circulating proteins related to CVD and inflammation (GP6, ST2, ALCAM) stratify a successful functional response to the REHAB-HF rehabilitation program in older adults following HF hospitalization. Further investigation is needed to identify proteins associated with a causal role for rehabilitation benefits.

## INTRODUCTION

Heart failure (HF) is a complex cardiovascular condition that often results in profound physical impairment such as frailty, poor exercise tolerance, limited mobility, and falls.^1^ These geriatric conditions, defined as multifactorial health conditions that occur in older adults and include frailty, mobility impairment, falls, and cognitive decline, are highly prevalent among older HF patients, resulting in prolonged recovery, reduced quality of life, and frequent hospitalizations.^2^ Guideline directed medical therapy, while essential for disease management, has limited impact on these geriatric impairments.^2^ In the Rehabilitation Therapy in Older Acute Heart Failure Patients (REHAB-HF) trial, a tailored, structured, and progressive multidomain physical rehabilitation intervention including strength, balance, mobility, and endurance delivered during and after hospitalization significantly improved 6-minute walk distance, frailty status, quality of life, and depressive symptoms compared with attention control, outcomes of particular importance for older adult with HF.^3^

Among older adults admitted with acute HF, there is a significant heterogeneity in the degree of physical impairment and functional decline after the HF hospitalization. Circulating protein biomarkers present a new approach to gain greater insights into pathophysiological mechanisms such as inflammation^4^, skeletal muscle metabolism^5,6^, and fibrosis^7^ that may affect recovery of physical function after HF admission. Circulating proteins are being investigated to predict incident HF^8,9^, differentiate HF phenotypes^10^, quantify severity of disease^11^, and potential as drug targets.^12^ Whether circulating proteins are effect measure modifiers (EMM) for treatment response or are mediators for the efficacy of physical rehabilitation HF hospitalization recovery remains unknown. In this investigation we aimed to: (1) identify and quantify if any specific circulating proteins can act as an EMM in the association between a physical rehabilitation intervention vs. attention control on functional recovery and clinical outcomes; (2) develop decision support for optimizing patient selection into a physical rehabilitation intervention; and (3) test whether changes in circulating protein mediate functional recovery, i.e. to extend effect modification (stratification) to a potential causal pathway.

## METHODS

### Data Availability

The data that support the findings of this study are available from the corresponding author upon reasonable request.

### Study Population

Details of the REHAB-HF trial (NCT02196038) design, intervention, and study populations have been previously published.^13,14^ Briefly, the study population consisted of 349 older adults ε60 years of age who were hospitalized with acute HF and exhibited at least one symptom and two signs of HF that resulted in a change to medical therapy targeting HF domains. While there was no exclusion criteria based on left ventricular ejection fraction (LVEF), participants had to have the ability to: (1) perform basic activities of daily living prior to their index HF admission; (2) walk independently with or without assistive device for at least four meters prior to enrollment; (4) be discharged to home.

### Proteomic Sub-Study Participants

All REHAB-HF participants were eligible for inclusion in this targeted discovery proteomics sub-study with availability of at least a baseline pre-randomization serum sample. The sub-study consisted of 243 of the 349 study participants with available baseline cryopreserved serum sufficient for proteomic analysis.

### Study Interventions and Outcomes

Participants were randomized to the rehabilitation intervention (RI) versus usual care attention control (AC). The RI consisted of a 12-week, 3 times per week tailored, structured, and progressive multidomain physical RI focusing on four domains necessary for independence - strength, balance, mobility and endurance - versus AC. Both arms could receive usual care medical and clinical therapies at the discretion of their treating physician. The primary outcome of the study was the change in the score of the Short Physical Performance Battery (SPPB), a composite test of leg strength, upright balance, and gait speed, from randomization (prior to hospital discharge) to 12-weeks. The SPPB is a 12-point ordinal scale, with higher scores indicating better physical function and reported in detail in the primary study results.^3^ A key goal of the study was to improve participant endurance measured by walking distance. Change in 6-minute walk distance (6MWD), measured in meters, was included as a co-primary endpoint in this sub-study of the Rehab-HF trial. As a continuous outcome, it offers a greater granularity than the SPPB and captures complementary physiological domains.^15^

### Proteomic Measurements

For the targeted discovery proteomic analysis, serum samples drawn at baseline pre-randomization and at 12-week follow-up were collected and stored at −80°C. Samples were sent to Olink (Watertown, MA) for analysis with the Olink Target-96 Cardiovascular III panel consisting of 92 unique proteins curated based on multiple knowledge based domains of cardiovascular disease measured using the proximity extension assay technology.^16^ Values are not reported as concentrations, but as normalized protein eXpression (NPX) values expressed in log base 2. This panel was selected based on its coverage of heterogeneous domains of cardiovascular disease pathways, prior work showing differential protein expression between patients with HFrEF and HFpEF, and our experience showing associations with functional parameters including 6MWD and clinical outcomes in clinical trial participants recently hospitalized with HF.^17^ Reproducibility and validation information regarding the proteins are reported by Olink.^16^ A list of proteins included in the analysis can be found at the Olink website.^16^ Quality control measures for the Olink Target-96 panels are available on the Olink website.^16^

### Statistical Analysis

Statistical analyses were performed using R statistical software version 4.3.1.^18^ Visualizations of variable distributions were developed to identify outliers and biologically implausible values. Four of the 92 proteins had some measurements below the level of detection (15% SPON 1, 4% CHIT1, 0.5% NT-proBNP, and 0.2% IGFBP-1). Rather than imputing or censoring these values, we retained instrument-reported measurements in our analysis.^18^ While these measurements are associated with increased uncertainty, this approach preserves the natural data distribution and avoids potential biases introduced by imputation and censoring.^19,20^ *Multi-faceted Approach to Discovering Effect Measure Modifier Proteins*

Effect modification resulting in stratification of treatment response can be characterized in multiple ways and may be affected by the presence of other variables, so we use a multi-faceted approach to test for effect modification under different model specifications (linear and non-linear), in the presence of different adjustment variables, and through different functional definitions for effect modification (traditional pairwise interactions and a tree-based characterization of interactions). First, we assessed individual linear regression models each including a single protein as an explanatory variable and adjusting for age, sex, race, HF subtype, baseline functional status (SPPB or 6MWD depending on outcome) and treatment assignment, with separate models for 12-week change in each of the two functional outcomes. To identify EMM proteins, interaction effects between treatment and baseline protein expression were assessed for significance using the Benjamini-Hochberg procedure^21^ to control the overall false discovery rate (FDR) at the 5% level. This one-protein-at-a-time marginal screening procedure may fail to identify EMM proteins that stratify treatment effect only when controlling for other proteins, so we next use a hierarchical LASSO procedure for simultaneous estimation and variable selection of EMM proteins in a single linear regression model.^22^ This model includes the same adjustment covariates but also adjusts for pairwise interactions among all proteins. Cross-validation is used to fine tuning parameters for the hierarchical Lasso such that the algorithm can identify significant protein-treatment interactions while enforcing strong hierarchy, a modeling principle that requires main effects for any two variables to be included in a model for which an interaction is also included between those variables. This avoids the detection of spurious interaction effects that are insignificant in the presence of the main effects. Lastly, we used the Bayesian additive regression tree (BART) method to identify EMM proteins that may stratify treatment effect in a nonlinear manner. BART has been shown to outperform competing models (LASSO, gradient boosting, neural nets, random forests) with respect to classification and estimation accuracy, accounts for complex interactions among proteins, and provides tools for variable selection and detecting effect modification.^23,24^ In this model, successive splits with a tree on treatment arm indicator and protein expression level represent interaction effects associated with effect measure modification. Importance scores can be constructed by counting the number of instances of such successive splits for each protein across all trees.

### Propensity Score Matching and Biomarker-based Classification and Regression Trees

To mitigate selection bias due to incomplete blood sample collection at baseline and to achieve marginal exchangeability between those randomized to the RI versus AC who had baseline blood samples for protein measurements, we identified matched pairs using the identified EMM proteins followed by demographic covariates and HF subtypes to achieve a 1 to 1 match of participants assigned to RI or AC with baseline biomarker measures.^25^ Then we used a classification and regression decision tree (CART) approach to model within-pair outcome differences to identify easily interpretable subgroup structure.^25,26^ CART models are extremely flexible and prone to overfit. Trees produced by CART were pruned using Least Squares Means simultaneous confidence intervals to prevent overfitting.^27^ For internal validation of the CART decision tree we used 10-fold cross-validation to identify the maximum tree depth and minimum number of observations in each terminal node that produced the lowest overall test set root mean square error (RMSE) separately for SPPB and 6MWD. We also compared the overall test set RMSE for the optimal decision tree based on cross-validation to that of a random forest model to compare relative model performance. For more details regarding our regression decision tree based on propensity score matching approach to model development, we have published the R code as a R Markdown file (**Supplementary Material – R-Code**).^28^

### Causal Mediation Analysis

In order to test for a possible indirect effect of treatment on improvement in functional outcomes mediated by 12-week change in EMM protein expression, we used a causal mediation analysis approach.^29–31^ For the identified EMM proteins, we first tested for differences in the 12-week change in protein levels across treatment groups using a nonparametric Wilcox test. Then, a linear regression framework was used to determine the proportion of the total effect of treatment on improvement in functional outcomes mediated by 12-week change in EMM protein expression representing the average causal mediation effect.

### Protein Clustering

As an exploratory analysis as an alternative to supervised machine learning approaches previously applied for the selection of single effect measure modifier proteins, we applied unsupervised learning with a weighted gene co-expression network analysis (WGCNA) approach to cluster proteins using baseline expression levels.^32^ WGCNA involves hierarchical clustering of proteins to form clusters of highly correlated proteins, which can then be summarized and tested for EMM accordingly.

### Methods Summary

A graphical summary of the study design with selection of participants, randomization to the intervention, functional outcomes integrating the selection of the EMM proteins, their use in a multi-marker approach to stratifying the intervention efficacy for functional outcomes and causal inferences assessment are encapsulated in the **Central Figure**. This analysis has been determined not to involve human subjects research as defined under 45 CFR 46.102(e) by the University of Maryland Baltimore Institutional Review Board.

## RESULTS

### Baseline Clinical Characteristics

A total of 243 of 349 (69.6%) of REHAB-HF participants had baseline proteomics measured. The mean age of the participants was 72.3±8.0 years, 52% were female, 55% of White race and 53% with HF with preserved LVEF. The median SPPB was 6.0 (Quartile 1[Q1] 4.0, Q3 8.0) and the median 6MWD was 183 (Q1 101, Q3 266) meters. In the 243 participants with serum available, demographic and clinical characteristics did not differ between participants randomly assigned to RI versus AC 12-week treatment arms of the study (**Table 1**).

**Table 1.**
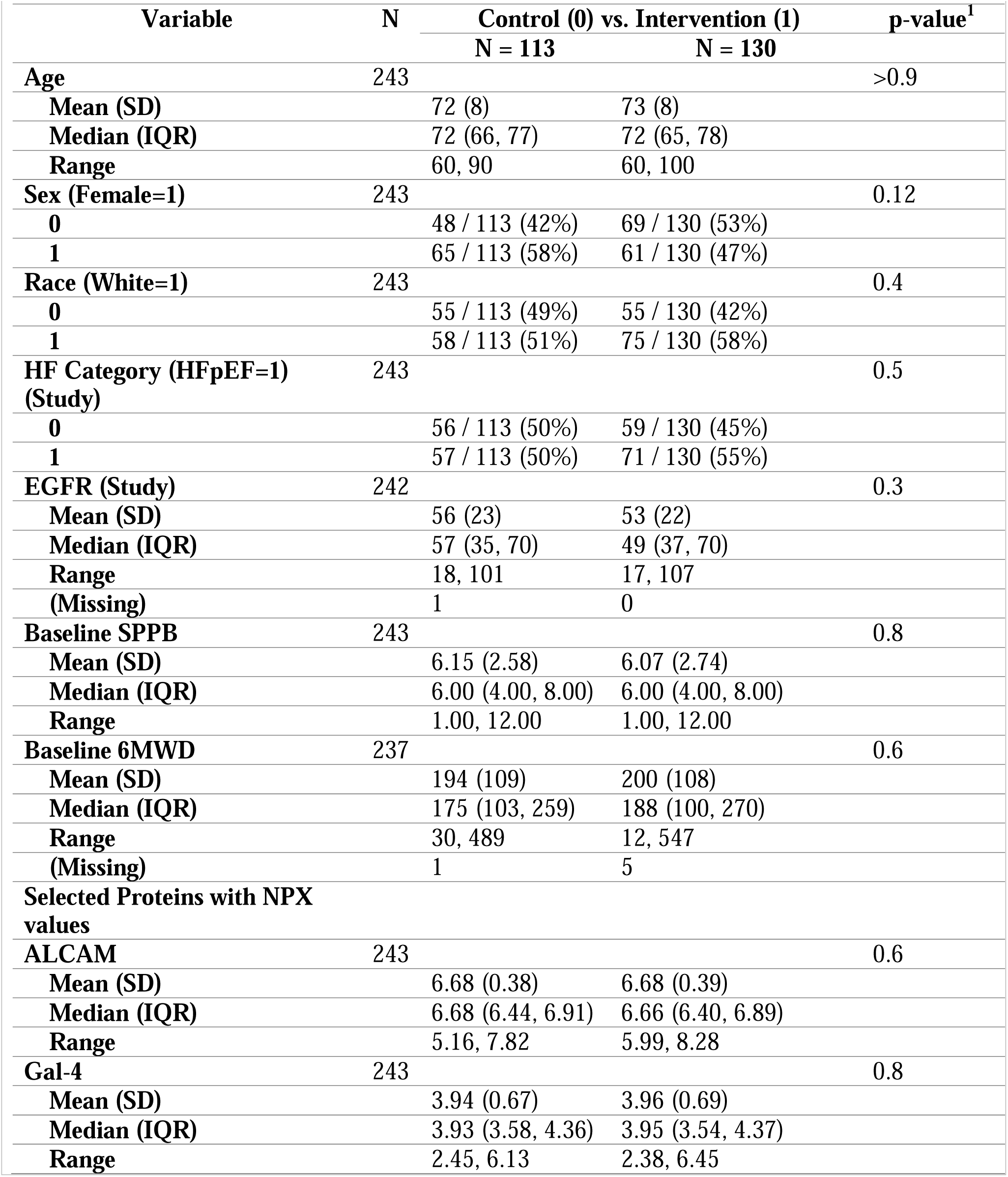
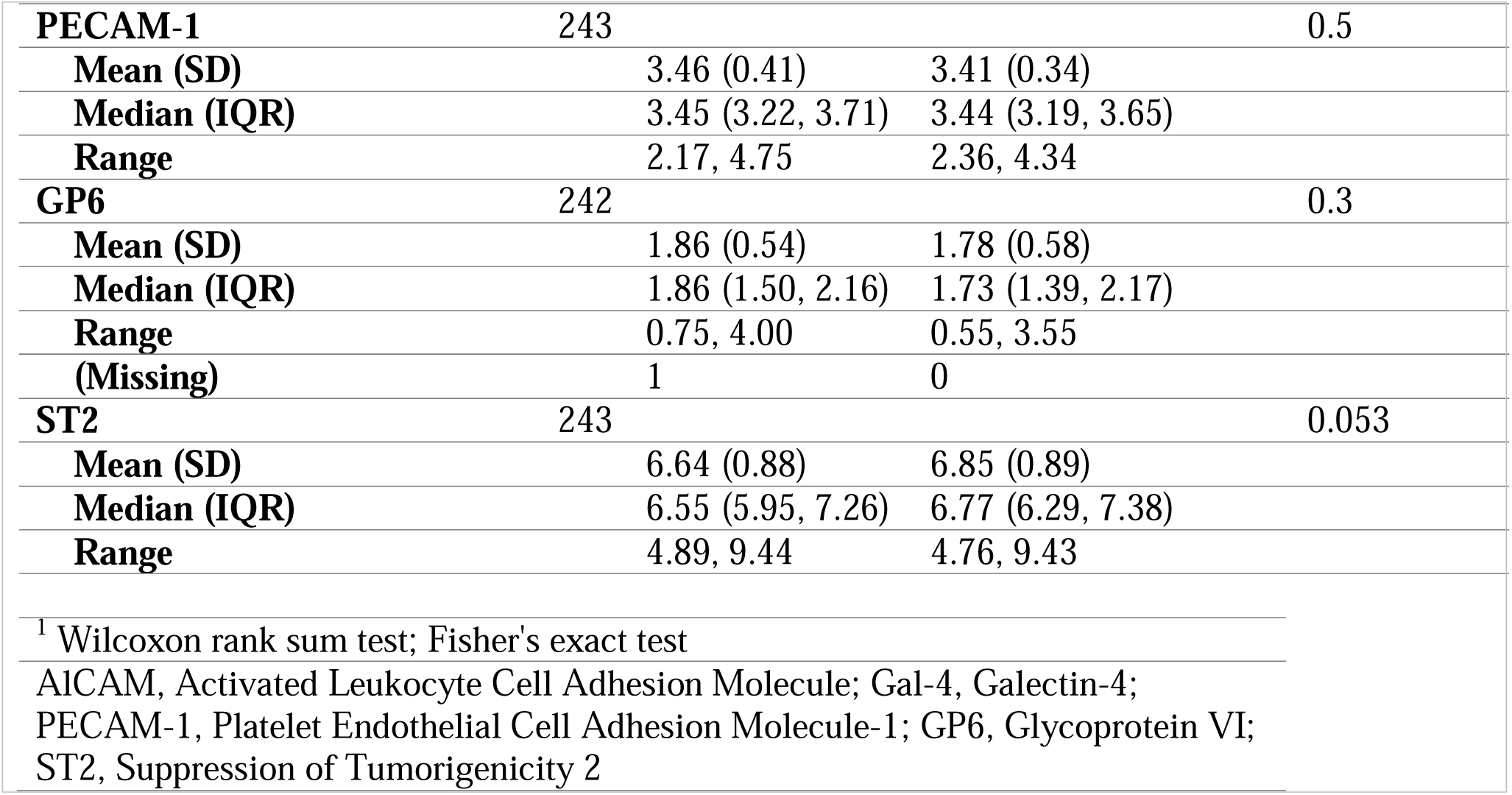
Baseline Patient Characteristics.

### Identification of Effect Measure Modifiers for SPPB and 6MWT

Participants with samples for baseline and follow-up measures as well as functional follow-up testing are shown in a consort diagram in **Supplemental Figure 1**. Of the 243 participants with baseline proteomic measures 213 (88%) returned in 12-weeks and had their SPPB assessed and 185 (76%) had their follow-up 6MWD measured. To select potential effect measure modifier (EMM) proteins and stratify the incremental improvement in both SPPB and 6MWD, we applied the three analytical approaches using the 92 circulating proteins available at baseline in all participants. First, multivariable linear regression models inclusive of a single protein as an explanatory variable identified 8 proteins with levels that had a nominal significant interaction (prior to correction for FDR) with treatment assignment for change in SPPB and 14 proteins that had a nominal significant interaction with treatment assignment for change in in 6MWD. However, all 8 and 14 proteins respectively no longer had statistically significant interactions with treatment assignment once adjusted for FDR. Therefore, based on this approach, no proteins were selected for additional EMM analysis.

Next, we applied hierarchical LASSO for protein selection and identified Platelet glycoprotein VI (GP6) as an EMM with a significant interaction with treatment assignment for SPPB and Activated Leukocyte Cell Adhesion Molecule (ALCAM) and Galectin-4 (Gal-4) as EMM proteins with a significant interaction with treatment assignment for 6MWD. Lastly, we used the BART method to relax the structure of the relationship between the protein expression levels, and the 12-week change in functional outcomes to no longer assume a linear relationship. Using this approach, we further selected based on visual inspection of importance plots (**Supplemental Figure 2)** soluble suppression of tumorigenicity 2 (ST2) for SPPB, and Gal-4 and Platelet Endothelial Cell Adhesion Molecule-1 (PECAM-1) for 6MWD. The correlations between baseline levels of the five proteins are shown as heatmap in **Figure 1**. ALCAM and PECAM-1 were significantly associated with all other proteins and most strongly correlated with each other. A modest, but significant correlation between ST2 and Gal-4 was also present.

**Figure 1.**
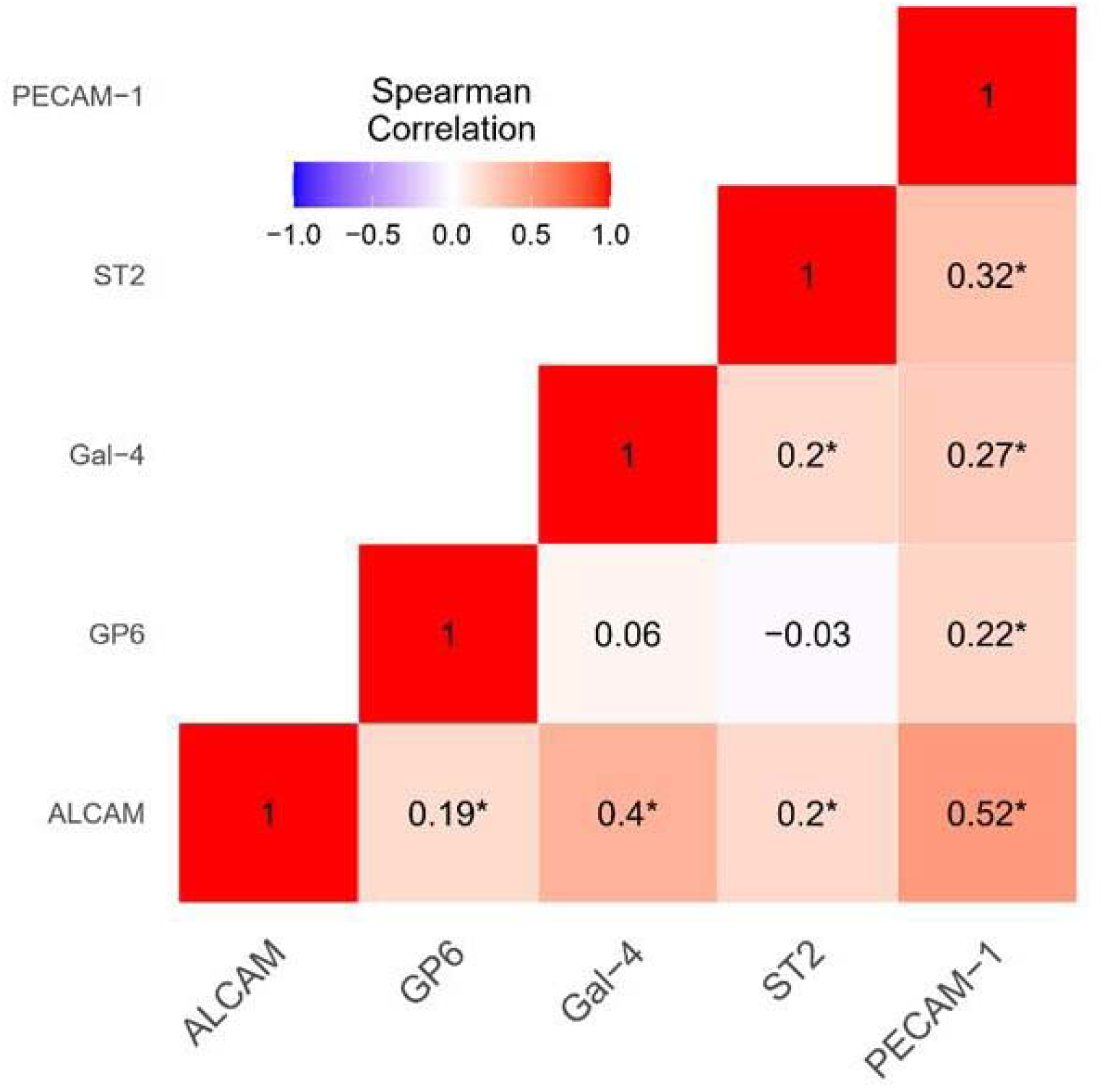
Correlation between baseline expression levels for five proteins selected by LASSO or Bayesian Additive Regression Trees as effect measure modifiers for the physical therapy rehabilitation intervention versus attention control treatment assignments for 12-week change in either functional outcome (SPPB or 6MWD). 6MWD, six-minute walk distance; ALCAM, Activated Leukocyte Cell Adhesion Molecule; Gal-4, Galectin-4; GP6, Platelet glycoprotein VI; PECAM-1, Platelet Endothelial Cell Adhesion Molecule-1; SPPB, Short Physical Performance Battery; ST2, soluble suppression of tumorigenicity 2.

Baseline expression levels in NPX units of the five selected EMM proteins are shown in Table 1. There is no difference in baseline levels based on treatment assignment.

### Effect Measure Modifiers: Functional Outcomes and Treatment Interaction

After identifying candidate EMM proteins, we examined their association with functional outcomes using marginal linear models to enhance interpretability and to assess interaction with treatment assignment. The overall mean SPPB at 12-weeks was 7.5±3.3 with a mean improvement from baseline of 1.2±2.8. The overall mean 6MWD at 12-weeks was 269±132 with a mean improvement from baseline of 66±94. The association with each of the previously selected EMM proteins with 12-week change in SPPB and 6MWD was evaluated. In **Table 2** each protein is shown as an independent variable per increase in NPX unit in unadjusted and adjusted linear regression models. An increase in one NPX unit is equivalent to a doubling in level. For 12-week adjusted change in SPPB higher baseline GP6 and ST2 expression levels are associated with a trend for less improvement (adjusted P values 0.051 and 0.059). Neither protein has a significant interaction with treatment assignment. For 12-week change in 6MWD, higher baseline expression levels of ALCAM and PECAM-1 after adjustment are associated with less improvement, estimated as a mean of −85.5 meters and −41.8 meters respectively for each increase in NPX unit. Both proteins have statistically significant interactions with treatment assignment.

**Table 2.** Associations of twelve-week change in functional outcomes with baseline protein levels by linear regression analysis.

| Protein | Unadjusted Effect | Unadjusted. P-value | Adjusted Effect | Adjusted P-value | Interaction P-value |
| --- | --- | --- | --- | --- | --- |
| **12-Week Change in SPPB (N=213)** |  |  |  |  |  |
| GP6 | -0.69 (-1.09, -0.30) | 0.001 | -0.56 (-1.12, 0.002) | 0.051 | 0.822 |
| ST2 | -0.76 (-1.67, 0.15) | 0.100 | -1.16 (-2.37, 0.05) | 0.059 | 0.093 |
| **12-Week Change in 6MWD (N=185)** |  |  |  |  |  |
| ALCAM | -13.19 (-51.67, 25.29) | 0.450 | -85.50 (-150.83, -20.17) | 0.011 | 0.013 |
| Gal-4 | 14.33 (-10.72, 39.37) | 0.261 | 31.56 (-12.70, 75.82) | 0.161 | 0.230 |
| PECAM-1 | -18.80 (-39.36, 1.77) | 0.073 | -41.75 (-72.11, -11.39) | 0.007 | 0.040 |

Adjustment covariates: age, sex, race, heart failure subtype (reduced or preserved ejection fraction) and treatment assignment. Interaction P-value is for protein level \* treatment assignment. Units for SPPB are based on an integer score of 0-12 and 6MWD is in meters. The effect size is per NPX expression unit of the protein which on a log2 scale such that an increase of one unit is equivalent to a doubling of the protein level. Abbreviations: SPPB: Short Physical Performance Battery; 6MWD: Six-Minute Walk Distance.

The change from baseline to 12-weeks for SPPB score and 6MWD stratified by baseline protein expression levels are visualized on a continuous scale separated by treatment assignment in **Figure 2** and **Figure 3**, respectively. For SPPB progressively higher levels of GP6 are associated with a trend for a greater improvement in SPPB when assigned to RI, but a decline in improvement for participants assigned to AC. In contrast, progressively higher levels of ST2 are associated with a seeming linear decline in improvement in SPPB score irrespective of treatment assignment. For 6MWD, increasing baseline levels of ALCAM, Gal-4 and PECAM-1 are associated with progressive separation in the 12-week improvement in 6MWD based on treatment assignment such that RI generally results in a greater 12-week improvement in 6MWD with higher baseline levels of the 3 proteins. In contrast, assignment to AC results in the opposite, with progressive decline in 12-week 6MWD improvement at higher baseline protein expression levels.

**Figure 2.**
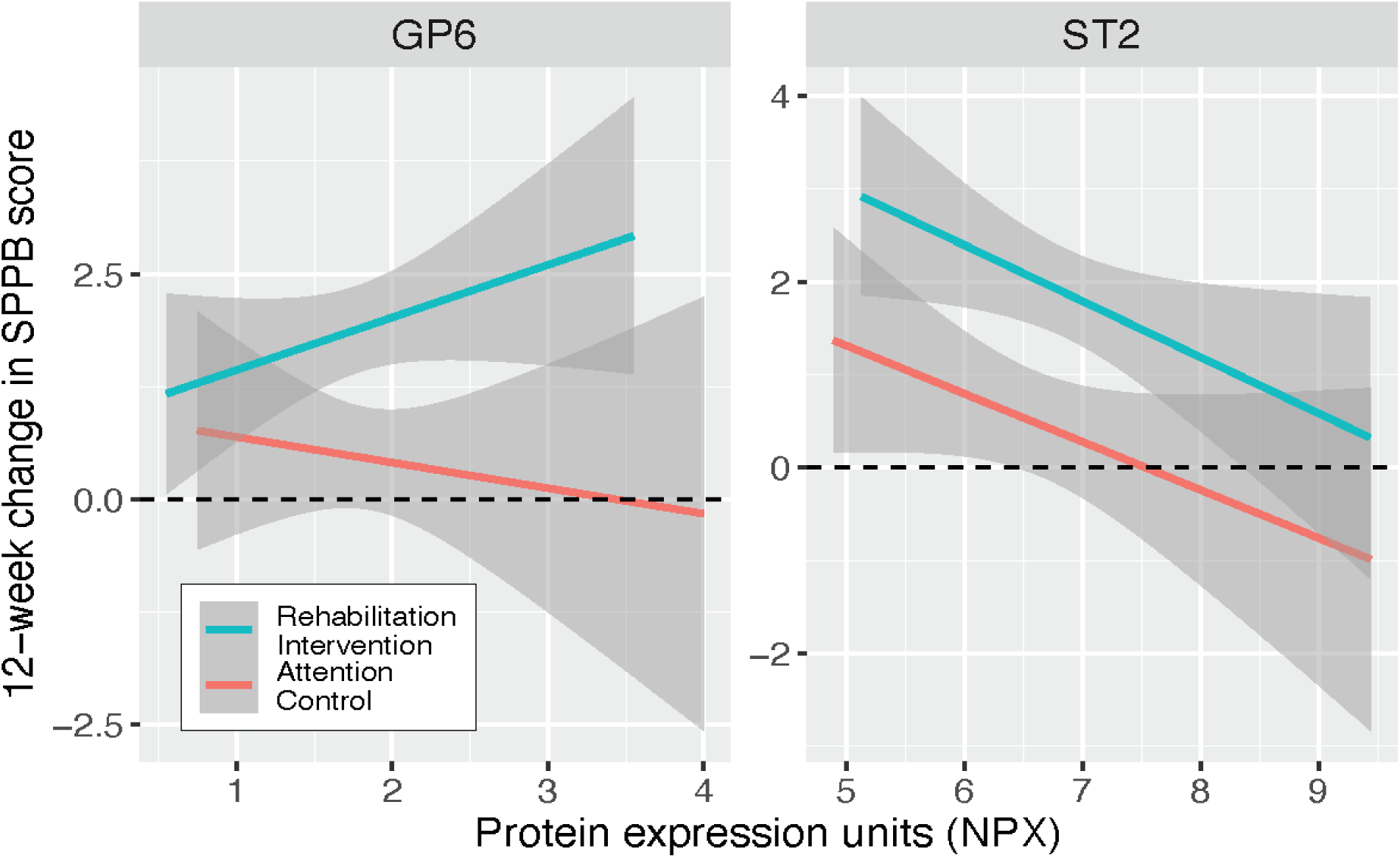
Linear associations between baseline protein expression levels and participants’ change from baseline to 12-weeks in the Short Physical Performance Battery differentiated based on the randomly assigned intervention (N=213 participants). GP6, Platelet glycoprotein VI; PECAM-1, SPPB, Short Physical Performance Battery; ST2, soluble suppression of tumorigenicity 2.

**Figure 3.**
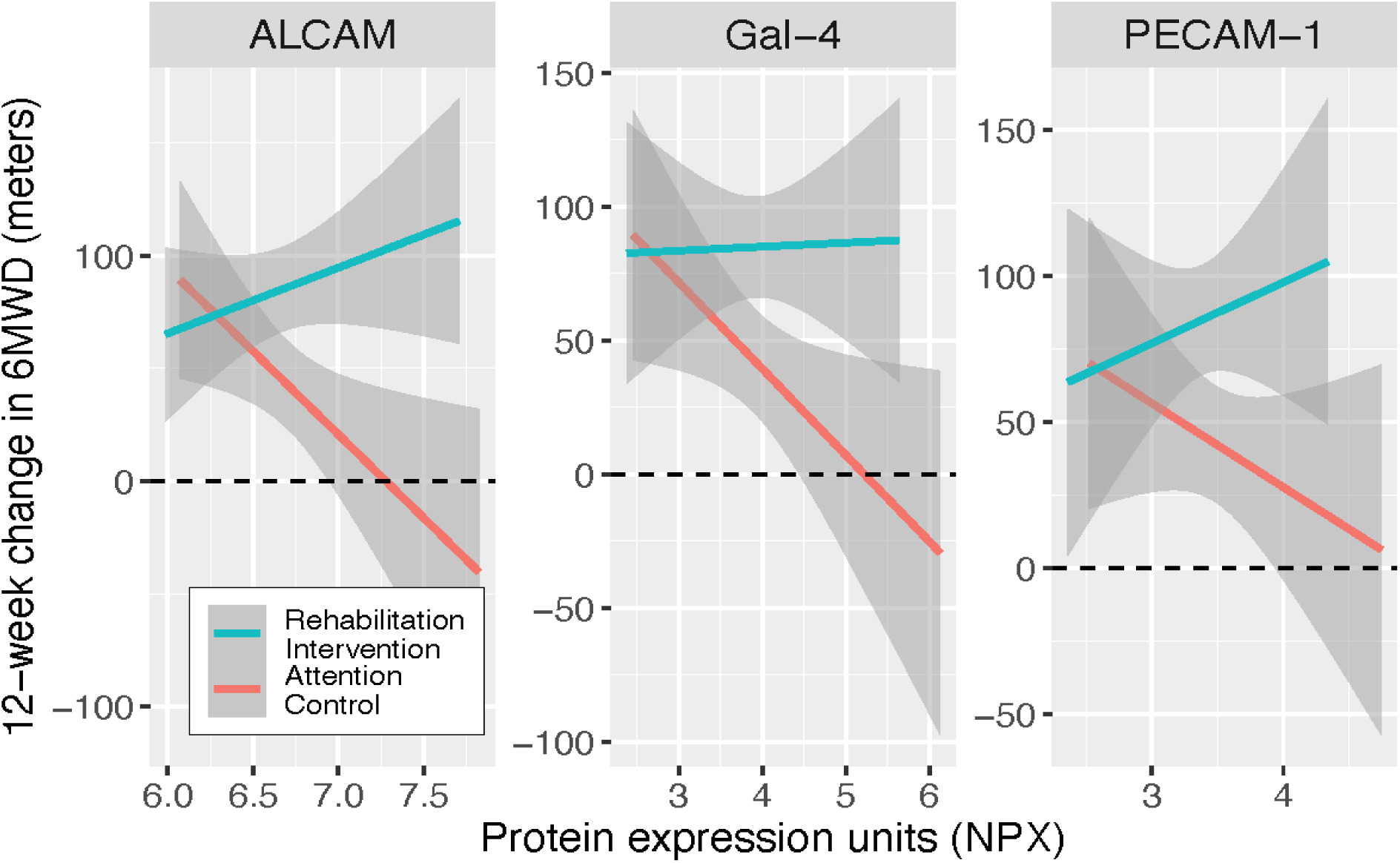
Linear associations between baseline protein expression levels and participants’ change from baseline to 12-weeks in six-minute walk distance differentiated based on the randomly assigned intervention (N=185 participants). 6MWD, six-minute walk distance; ALCAM, Activated Leukocyte Cell Adhesion Molecule; Gal-4, Galectin-4; GP6, PECAM-1, Platelet Endothelial Cell Adhesion Molecule-1; ST2, soluble suppression of tumorigenicity 2.

### Decision Trees for a Multi Protein approach to Stratification of Functional Outcomes Based on Treatment Assignment

To estimate optimal decision-points for previously selected individual EMM proteins when used together to stratify the differential change in SPPB and 6MWD from baseline to 12 weeks based on treatment assignment, we evaluated CART with prior propensity matching pairs of participants assigned to RI vs AC who had both outcomes measured at baseline and 12-weeks (**Supplemental Figure 1**). Propensity matching resulted in 162 (89.5%) and 142 (78.5%) of the 181 participants, with both SPPB and 6MWD measured at baseline and 12-weeks and baseline proteomic measures, being matched 1 RI participant to 1 AC participant for the outcomes of SPPB and 6MWD, respectively. Using these propensity score matched participants; we developed separate decision trees for SPPB and 6MWD with the unique EMM proteins selected for each functional outcome. The optimized cross validated decision trees compared favorably to a random forest model (**Supplemental Figure 3**). The decision trees for each functional outcome are shown in **Figure 4**. For SPPB, the presence of a baseline ST2 NPX expression level ε 6.3 measured in a participant is associated with the greatest differential improvement such that a participant randomly assigned to RI had on average an SPPB score that improved by 2.1 (95% CI 1.5, 2.7) points more than a participant assigned to AC. In contrast, for a participant with an ST2 < 6.3 NPX units and a GP6 < 1.9 NPX there was no difference in change in SPPB scores based on treatment assignment (**Figure 4a**). For 6MWD, participants were optimally differentiated based on ALCAM expression alone, such that participants with an ALCAM ε 6.7 NPX expression units assigned to RI had on average an 85 meter (95% CI 56m, 113m) greater increase in walking distance versus participants assigned to AC. In contrast, participants with an ALCAM < 6.7 NPX expression units had no difference in 6MWD at 12-weeks based on treatment assignment (**Figure 4b**).

**Figure 4.**
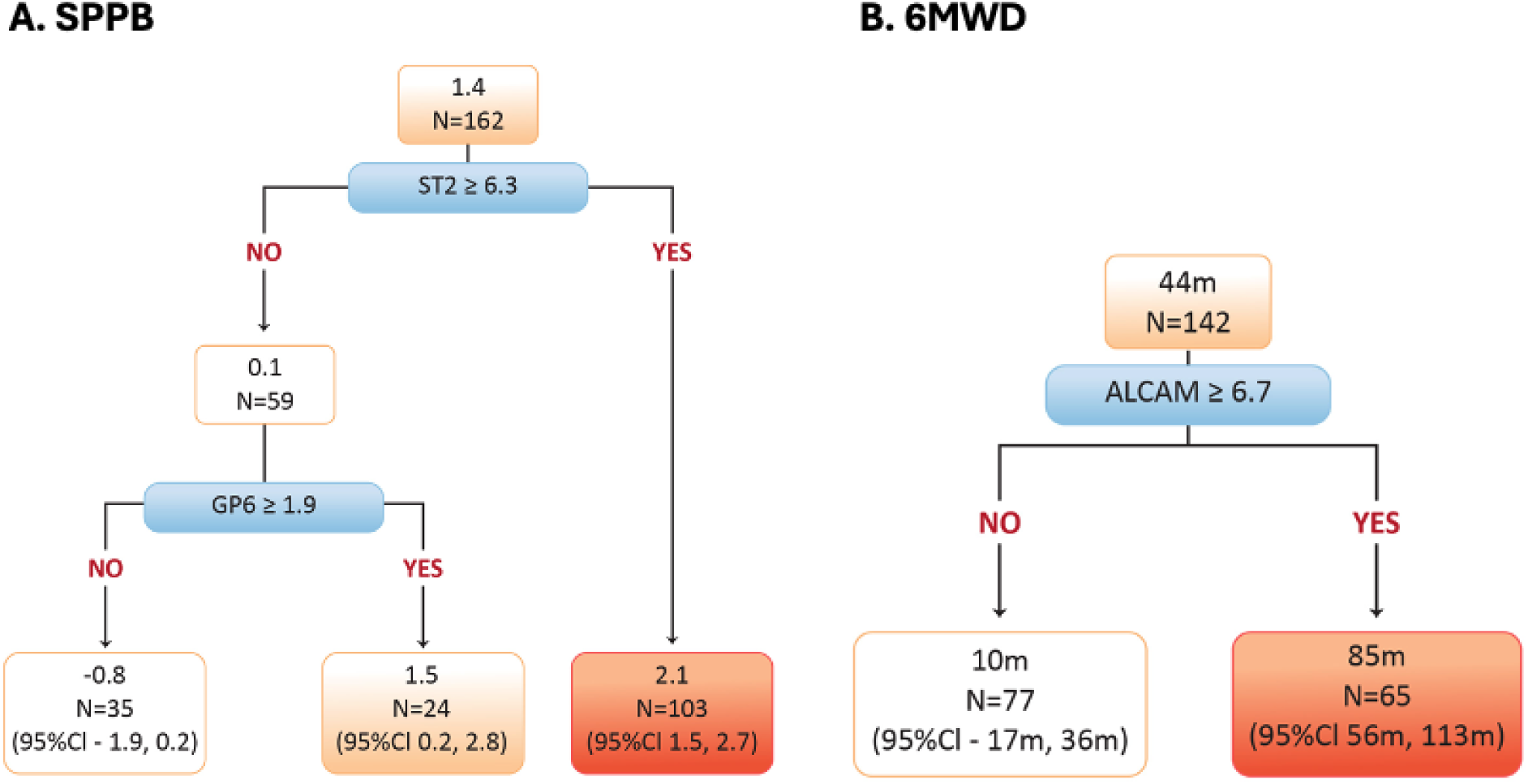
Classification and regression trees for the differential 12-week functional outcomes SPPB (panel A) and 6MWD (panel B) based on treatment assignment to the physical therapy rehabilitation intervention versus attention control using previously identified effect measure modifier proteins selected for each functional outcome. A positive number for 12-week change favors the rehabilitation intervention. Numbers shown are the difference in the 12-week functional outcomes between the two treatment assignments. For example, in panel B, in the right lower leaf, participants assigned to the physical therapy rehabilitation intervention had, on average, an 85-meter greater increase in walk distance from baseline to 12-weeks compared to participants assigned to attention control. 6MWD, six-minute walk distance; ALCAM, Activated Leukocyte Cell Adhesion Molecule; Gal-4, Galectin-4; GP6, Platelet glycoprotein VI; PECAM-1, Platelet Endothelial Cell Adhesion Molecule-1; SPPB, Short Physical Performance Battery; ST2, soluble suppression of tumorigenicity 2.

### Twelve-Week Change in Protein Levels and Mediation of Functional Outcomes

The change in each of the five proteins stratified based on treatment assignment to the RI versus AC is shown in the box plots in **Supplemental Figure 4**. Four of 5 proteins’ expression levels significantly changed from baseline to 12-weeks including a decrease in GP6 and ST2, and an increase in ALCAM and Gal-4. None of the 5 proteins’ 12-week change in expression level was different based on treatment assignment. Next, we evaluated whether changes in any of the five EMM proteins mediated the efficacy of RI for the two outcomes, SPPB and 6MWD. Specifically, we assessed the proportion of the treatment effect on improvement in these outcomes that was mediated by changes in protein levels (indirect effects, represented by ACME [Average Causal Mediation Effect]) versus the proportion attributed to the intervention independent of protein changes (direct effects, represented by ADE [Average Direct Effect]). We found no significant ACME for SPPB or 6MWD 12-week change from any of the five proteins on the differential treatment efficacy of the RI versus AC. **(Supplemental figures 5)**

### Protein Cluster Association with Baseline Characteristics and Functional Outcomes

As an exploratory analysis we applied unsupervised learning with WGCNA to cluster proteins using their baseline expression levels. We identified five unique clusters with hub proteins led by Matrix metalloproteinase-9 (MMP-9), Neurogenic locus notch homolog protein 3 (Notch-3), Insulin-like growth factor-binding protein 2 (IGFBP-2), Cystatin-B (CSTB), Ephrin type-B receptor 4 (EPHB4). Cluster membership ranged from 8-35 proteins. Each module (cluster) was assigned a color name and the hub protein and top four cluster proteins determined by descending eigengene values are shown in **Supplemental Table 1**. Two clusters, brown (hub protein CSTB) and yellow (hub protein IGFBP-2) were positively associated with age. No clusters were associated with sex. All 5 clusters were inversely associated with baseline 6MWD and 3 of 5 clusters were inversely correlated with baseline SPPB. No clusters were associated with 12-week change in SPPB or 6MWD (**Supplemental Figure 6**). In a linear regression analysis adjusting for age, sex, HF subtype and intervention assignment, none of the 5 clusters had a significant interaction with treatment and therefore were not considered effect measure modifiers.

## DISCUSSION

The REHAB-HF trial previously demonstrated that the multidomain physical rehabilitation intervention significantly improved physical function capacity and quality of life in older adults hospitalized for acute heart failure with geriatric syndromes compared to attention control. In this current investigation, we extended these findings by exploring the role of circulating proteins as effect measure modifiers to identify patients most likely to benefit from RI. Using traditional statistics (linear regression), none of the 92 cardiovascular related proteins assayed were statistically significant effect measure modifiers of the rehabilitation effect on 12-week physical function after controlling for multiple testing. However, using complementary machine learning selection methods (hierarchical LASSO regression and Bayesian additive regression trees), we found that baseline levels of several proteins, specifically platelet glycoprotein VI (GP6), soluble suppression of tumorigenicity 2 (ST2), activated leukocyte cell adhesion molecule (ALCAM), and platelet endothelial cell adhesion molecule-1 (PECAM-1) significantly modified the treatment effect of the RI on functional outcomes.

Higher baseline levels of selected proteins were associated with differential responses to RI versus AC. For example, patients with elevated ALCAM or PECAM-1 expression levels at baseline experienced greater improvements in 6MWD with RI compared to those with lower levels, suggesting certain indicators may identify super-responders to the physical therapy rehabilitation intervention. Participants with higher baseline ALCAM (ε6.7 NPX units) assigned to RI had substantially greater improvements in 6MWD, approximately 85 meters, compared to AC, where differential improvement was minimal in those with lower baseline ALCAM.

Overall, higher baseline soluble ST2 expression levels were associated with attenuated improvements in SPPB scores regardless of treatment assignment. This has similarities to other post HF rehabilitation studies. For example, in the HF-ACTION trial of traditional aerobic exercise training (distinct from the multidomain RI in REHAB-HF) in chronic HF with reduced ejection fraction (distinct from acute decompensated HF regardless of ejection fraction in REHAB-HF), lower baseline soluble ST2 levels were associated with great survival benefit from exercise, whereas patients with very high ST2 levels experienced higher mortality risk.^33^ In our older adult post-hospitalization HF cohort, we observed a somewhat different pattern on functional outcomes. Patients with elevated ST2 levels (a marker of cardiac stress, type 2 inflammation, and fibrosis) generally experienced less improvement in SPPB score, but RI recipients achieved absolute gains in SPPB nonetheless. In contrast, those with higher ST2 levels in the AC arm tended to have a decline in SPPB 12-weeks post hospitalization. This suggests that RI may be able to offset the deficit in functional recovery associated with a high-risk immunologic profile. ST2 is an established risk factor in patients with HF because it is strongly associated with hospitalization and mortality risk.^34^ Our findings extend this knowledge by exploring ST2 in relation to functional outcomes. While ST2 did not show interaction when evaluated with traditional linear regression statistics, a decision tree analysis that does not assume a linear relationship between explanatory features (i.e., the protein levels) and functional outcomes indicated that patients with a higher relative ST2 threshold derived clinically meaningful improvement (+2 points) with RI versus AC that was not observed in participants below that threshold. ST2 may be a marker for a frailer high risk older patients with underlying immunologic dysfunction who particularly may need and respond to RI.

In this study, the adhesion molecules involved in endothelial and immune cell interactions, ALCAM and PECAM-1, were identified as effect measure modifiers for the endurance outcomes 6MWD. A higher baseline ALCAM threshold in the decision tree analysis, was associated with more than 85-meter greater improvement in 6MWD with RI compared to AC. Participants with ALCAM less than this threshold did not achieve the same benefit. Higher levels of ALCAM are known to be elevated in patients with higher inflammatory burden and are associated with adverse cardiovascular events. In 5,165 patients with acute coronary syndromes, ALCAM was found to be an independent risk factor for with cardiovascular death.^35^ ALCAM is involved in T lymphocyte activation by stabilizing the immunologic synapse and may also promote monocyte transmigration across in an activated endothelium.^36^ In recently hospitalized HF populations, ALCAM may capture a high risk immuno-phenotype that predisposes to subsequent physical and functional decline and signifies a greater capacity for improvement if RI is applied.^37^

A similar hypothesis applies to PECAM-1 (CD31), which is also an adhesion molecule that reflects endothelial activation and leucocyte transmigration.^38^ In our study, PECAM-1 (CD31) was associated with poor 6MWD outcomes in participants in the AC group, but substantial improvement with RI. Exercise has been reported to reduce levels of other soluble adhesion molecules such as VCAM-1 and ICAM-1 and improve vascular function in patients with cardiovascular disease.^39^ Thus, we hypothesize that patients with higher endothelial activation and inflammatory burden may benefit from tailored multidomain physical rehabilitation interventions because of improvement of peripheral perfusion, endothelial cell reprogramming, muscle function, and overall endurance or physical capacity.

Platelet glycoprotein VI (GP6) was an effect measure modifier for improvement in SPPB but exhibited some complex patterns. Higher baseline GP6 tended to correlate with less improvement in SPPB overall, but there was an interaction that mitigated this effect in the RI group compared to usual care. GP6 is a platelet collagen receptor that plays a role in thrombosis and inflammatory processes.^40^ Higher GP6 could indicate a prothrombotic and proinflammatory state that impairs skeletal muscle perfusion and/or recovery. In our analysis, those patients with higher GP6 levels who received AC had notable poor functional gains, but those who had RI had modest improvement in function. Finally, Galectin-4 was also identified by the LASSO and BART methodologies on the 6MWD 12-week response to RI. Galectin-4 is implicated in metabolic inflammation and is found to be elevated in conditions like obesity and diabetes mellitus, atherosclerosis, HF, and mortality.^41^

This study carries potential research and clinical implications by pointing to a more personalized approach to a rehabilitation intervention in recovery from acute decompensated HF hospitalization. Measuring these proteins could help predict potential participants who would most likely benefit from the intervention. A simple decision tree with single cut-points for ST2 or ALCAM could serve as a starting point for risk stratification. Patients above thresholds for these proteins may derive clinically meaningful improvements in physical function. These findings highlight that chronological age alone should not exclude older recently hospitalized HF patients from rehabilitation; instead, a multimodal assessment could better identify those who respond best to therapy. Moving forward, integrating proteomic or other biomarker data with clinical factors may enable personalized rehabilitation strategies, for example designing more intensive programs based on threshold criteria for acute HF patients.

It is important to discuss what this study did not find. First, none of the baseline protein clusters, derived from network analysis, showed interaction with treatment. Thus, no single broad physiological domain of statistically interrelated proteins clearly stratified outcomes for RI. Instead, the signals of effect modification were protein specific. Second, we did not find evidence that changes in these individual proteins mediated the benefit of RI. While four proteins showed significant changes from baseline to 12 weeks, none of them differentially changed by treatment assignment. Thus, improvements in physical function achieved by the RI were not explained by changes in circulating protein levels. This suggests that these proteins are markers that correlate with patient phenotype (both frailty and resilience), rather than part of the causal pathways of RI efficacy. For example, patients with high ALCAM or ST2 may have naturally worse recovery, but they can overcome these physical deficits with a comprehensive RI. These proteins act as risk stratifiers and not a therapeutic target for functional recovery.

This study has several potential methodological limitations. First, this was a secondary observational analysis within a clinical trial (n=243 with proteomic data). Using traditional linear regression analysis, there was no statistically significant interaction between proteins and treatment assignment after adjusting for multiple comparisons. To attempt to mitigate this, we used robust feature selection and cross validation (e.g., LASSO, BART, and CART methods) with machine learning to converge on a consistent set of proteins. Second, the protein panel was limited to 92 preselected cardiovascular related proteins. It is probable that additional important EMM proteins exist outside this panel. Third, the proteins were measured using expression units and we can’t directly convert this to a specific concentration unit to suggest as a threshold.

Orthogonal testing with ELISA will be needed to assign concentration thresholds. Finally, there is inherent complexity in interpreting multiple biomarkers that are inter-correlated. In our data, ALCAM and PECAM-1 were moderately correlated and ST2 and Galectin-4 also showed a correlation. It is likely that these biomarkers point towards a common underlying patient phenotype. Future research is needed to confirm these findings and determine whether incorporating proteomic assessment into a multidomain physical rehabilitation intervention referral can improve outcomes for older adults with heart failure.

## CONCLUSION

In the REHAB-HF trial, using a pipeline based on traditional statistical and machine learning approaches, we report several candidate proteins that could stratify the effect of a multidomain physical rehabilitation intervention on functional recovery. These proteins are largely linked to aging, immune dysregulation, platelet activation, and endothelial health. While RI benefited on average the older adult admitted with acute HF, those with baseline high circulating expression levels of ALCAM, ST2, and GP6 were likely to derive larger functional benefits versus AC. Measurable circulating proteins representing multiple biological factors may be used to identify a cohort of older patients at higher risk that may benefit from a more intensive multidomain physical rehabilitation program.

## ACKOWLEDGEMENTS

The authors thank Devon Stuart for her creation of the central figure.

## FUNDING SUPPORT

Supported by an NIH Career Development Award [K23HL153771] to A.A.D and partially funded by institutional support from the Inova Health System Foundation.

## Supplemental Table and Figures

**Supplemental Table 1.** WGCNA clusters (modules) with hub and top additional 4 proteins.

|  | Top Proteins |  |  |  |  |  |
| --- | --- | --- | --- | --- | --- | --- |
| Module | Number of Proteins | 1 (Hub) | 2 | 3 | 4 | 5 |
| Green | 8 | MMP-9 | AZU1 | PRTN3 | MPO | PAI |
| Blue | 21 | Notch 3 | IGFBP-7 | ALCAM | CDH5 | IL-1RT1 |
| Yellow | 10 | IGFBP-2 | OPN | AXL | SHPS-1 | IGFBP-1 |
| Brown | 13 | CSTB | U-PAR | JAM-A | TFF3 | FABP4 |
| Turquoise | 35 | EPHB4 | TNF-R1 | LTBR | TNFRSF14 | IL-18BP |

Abbreviations for hub proteins. Other full protein names can be found at
https://olink.com/biomarker-search. CSTB, Cystatin-B; EPHB4, Ephrin type-B receptor 4; IGFBP-2, Insulin-like growth factor-binding protein 2; MMP-9, Matrix metalloproteinase-9; Notch-3, Neurogenic locus notch homolog protein 3.

**Supplemental Figure 1.**
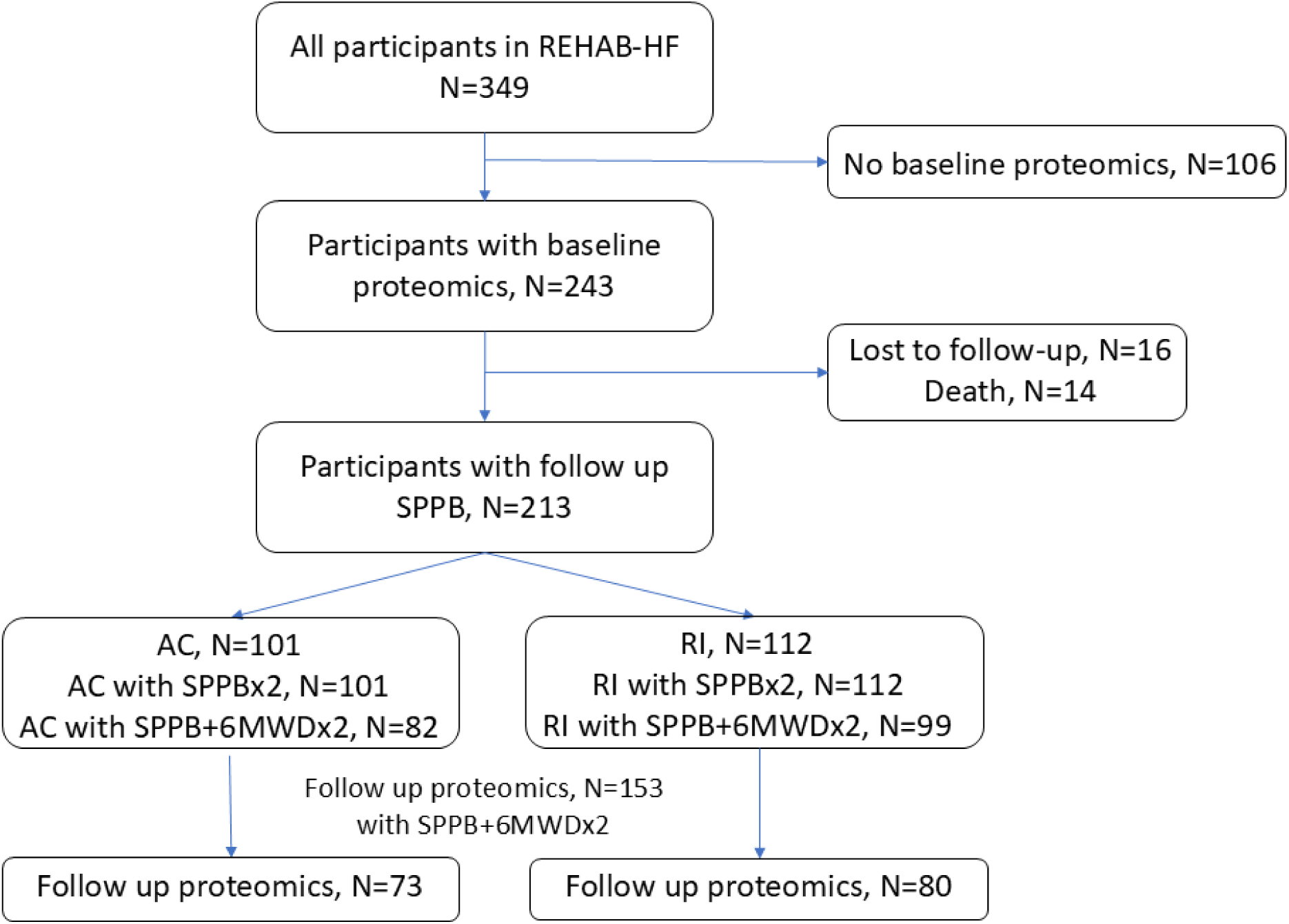
Consort diagram of participants in REHAB-HF who had baseline and follow-up proteomics measured along with the number with follow-up proteomic measures who also had SPPB and 6MWD performed. Note that 185 participants with baseline proteomics had a 6MWD measure at baseline and 12-weeks. Four of these participants did not have a follow-up SPPB reflected in the AC+RI combined groups with 181 participants with both SPPB and 6MWD at baseline and follow-up.

**Supplemental Figure 2.**
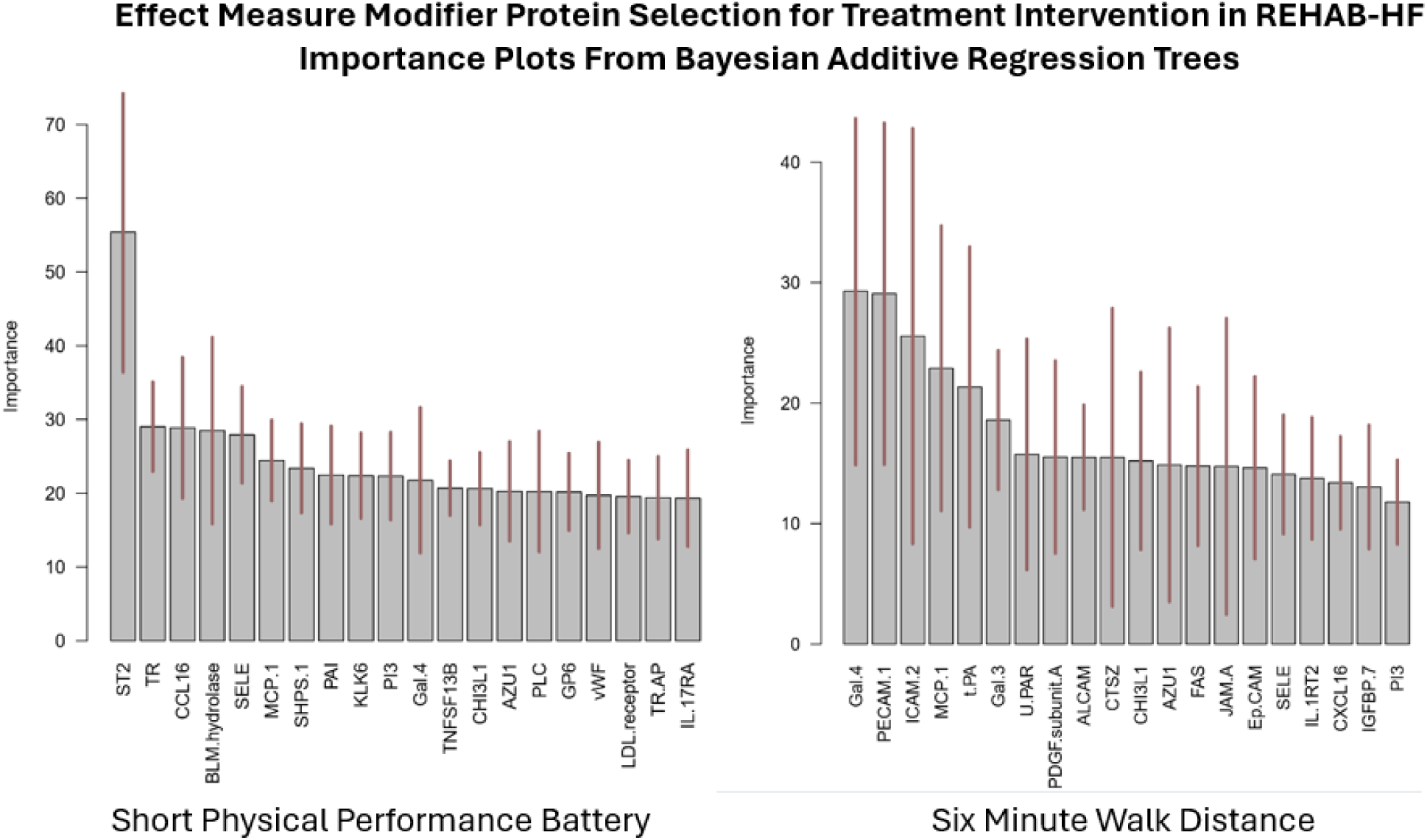
Importance plots using Bayesian Additive Regression Trees for the selection of baseline protein levels as effect measure modifiers of the differential 12-week response to the rehabilitation intervention versus attention control. Higher importance number indicates a larger role for a specific protein as an effect measure modifier for the differential efficacy of the intervention.

**Supplemental Figure 3.**
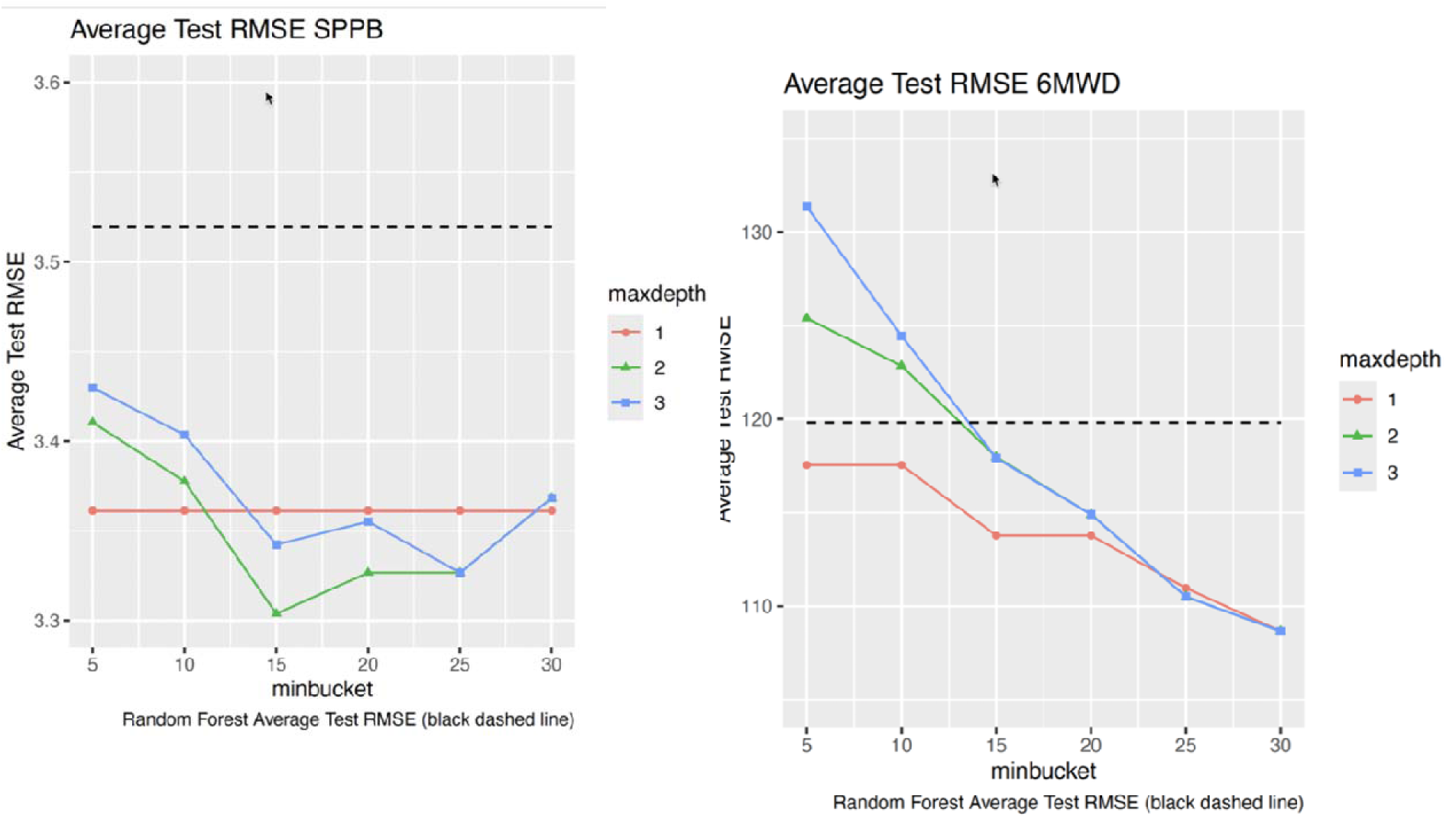
The 10-fold cross validation test set root mean square error (RMSE) for the classification and regression tree models are shown and optimized for minimal number of participants is any node (minbucket, x-axis) and maximal number of levels (maxdepth, colored lines) for SPPB (left panel) and 6MWD (right panel). The optimized decision trees had lower RMSE than Random Forest (see methods reference for Random Forest settings).^28^

**Supplemental figure 4.**
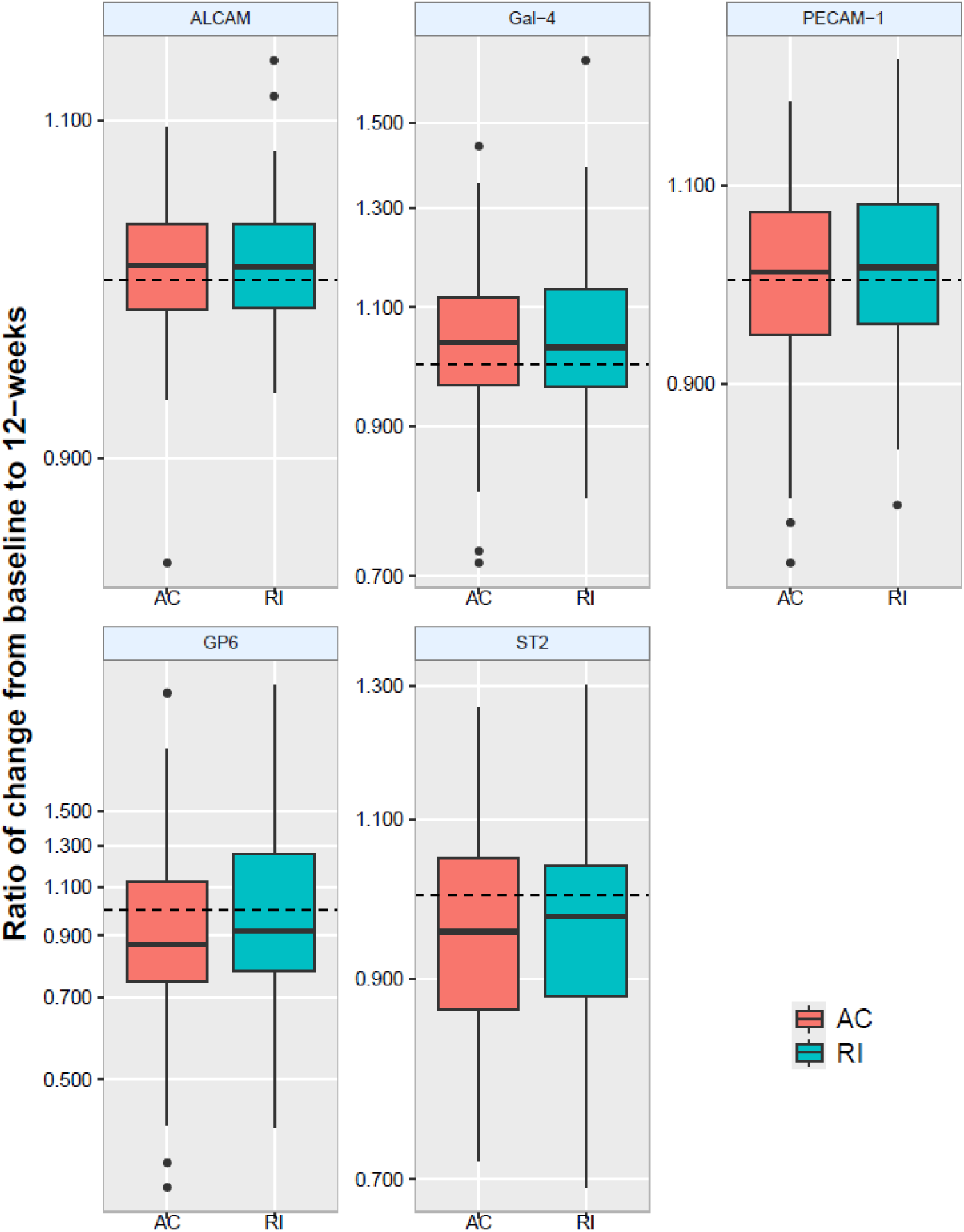
Change in circulating protein expression level from baseline to 12-weeks (n=155) for the five effect measure modifier proteins shown as the ratio to the baseline levels. Dark horizontal line is the median, and bottom at top edges of the boxes are the 25th and 75th percentiles respectively. No proteins’ change was different based on intervention assignment. GP6 and ST2 NPX expression levels significantly decreased and ALCAM and Gal-4 significantly increased from baseline to 12-weeks. AC, Attention control; ALCAM, Activated Leukocyte Cell Adhesion Molecule; Gal-4, Galectin-4; GP6, Platelet glycoprotein VI; PECAM-1, Platelet Endothelial Cell Adhesion Molecule-1; RI, rehabilitation intervention; ST2, soluble suppression of tumorigenicity 2.

**Supplemental figure 5.**
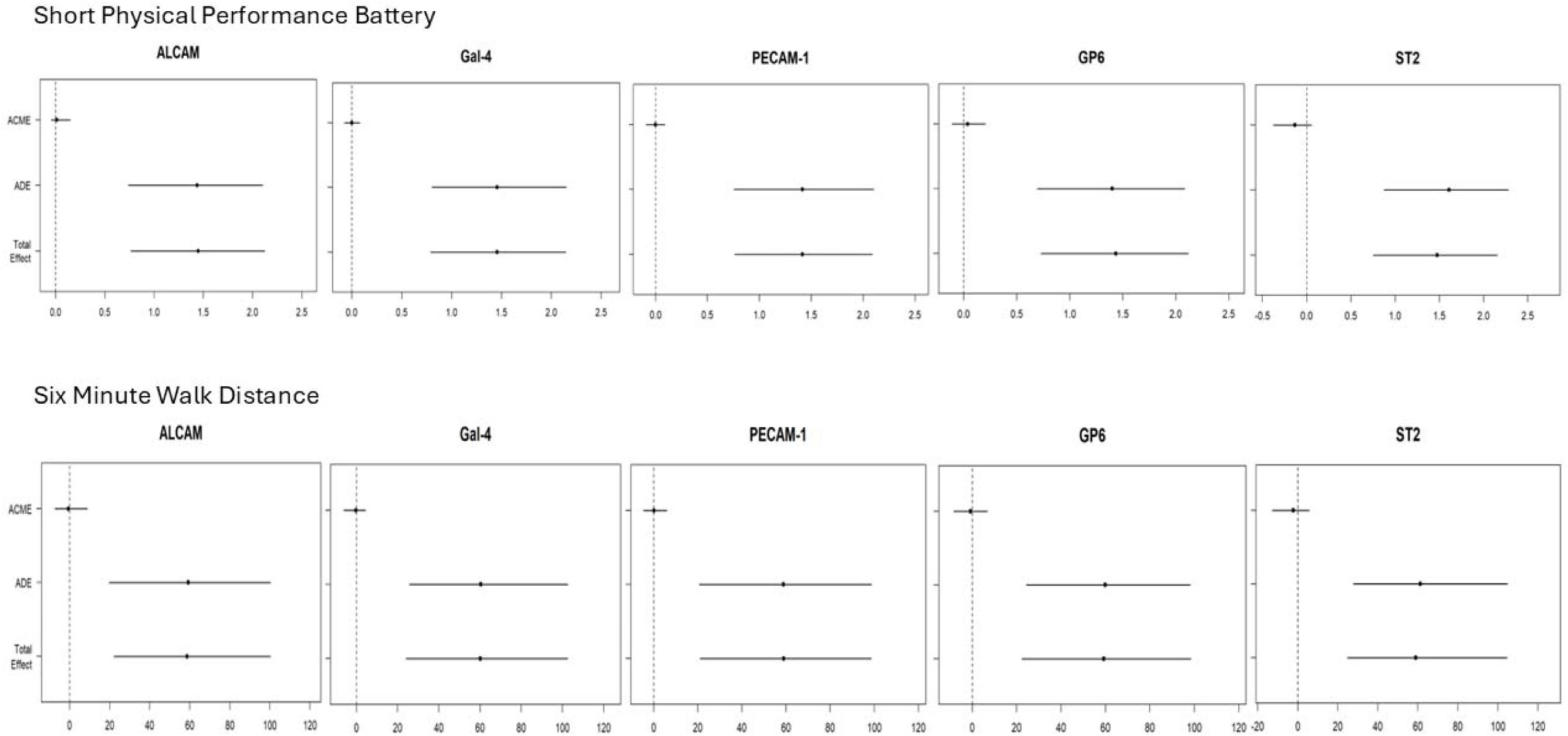
The mediation analysis shows the 12-week differential improvement in SPPB and 6MWD for the rehabilitation intervention versus attention control that are mediated through the change in protein expression levels as the Average Causal Mediation Effect (ACME) and the change not mediated through the change in protein expression level as the Average Direct Effect (ADE). The 12-week change in protein levels was not a mediator for the change in either functional outcome. Horizontal bars represent 95% confidence intervals.

**Supplemental figure 6.**
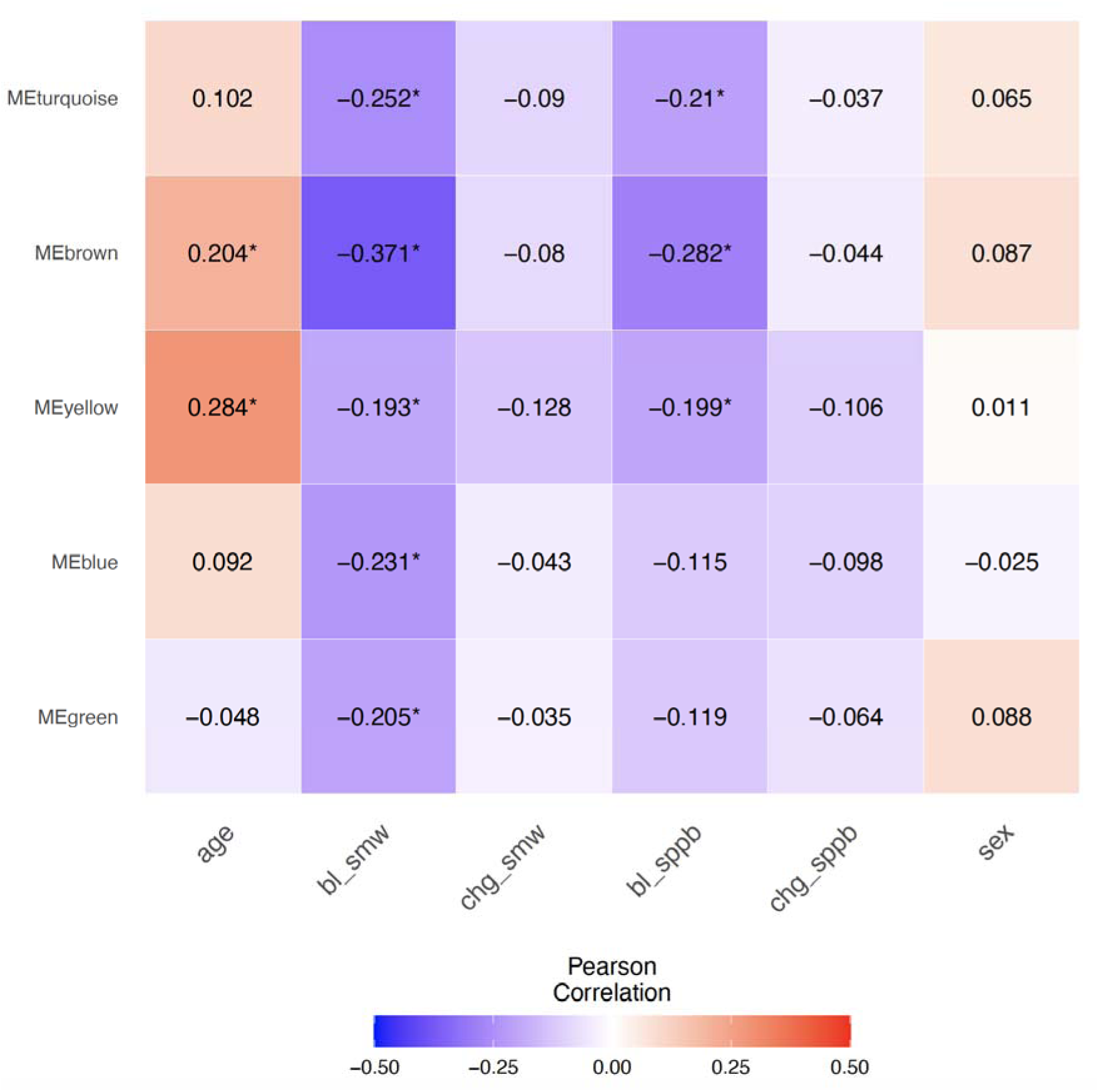
Correlation heat map of the five WGCNA protein clusters with age, baseline and 12-week change in functional outcomes. SMW, six-minute walk; SPPB, short physical performance battery, and sex. Clusters are labeled with a color and the hub protein assigned in supplemental table 6. Abbreviations: CSTB, 6MWD, six-minute walk distance, Cystatin-B; EPHB4, Ephrin type-B receptor 4; IGFBP-2, Insulin-like growth factor-binding protein 2; MMP-9, Matrix metalloproteinase-9; Notch-3, Neurogenic locus notch homolog protein 3; SPPB, short physical performance battery.

---

**Central Figure.**
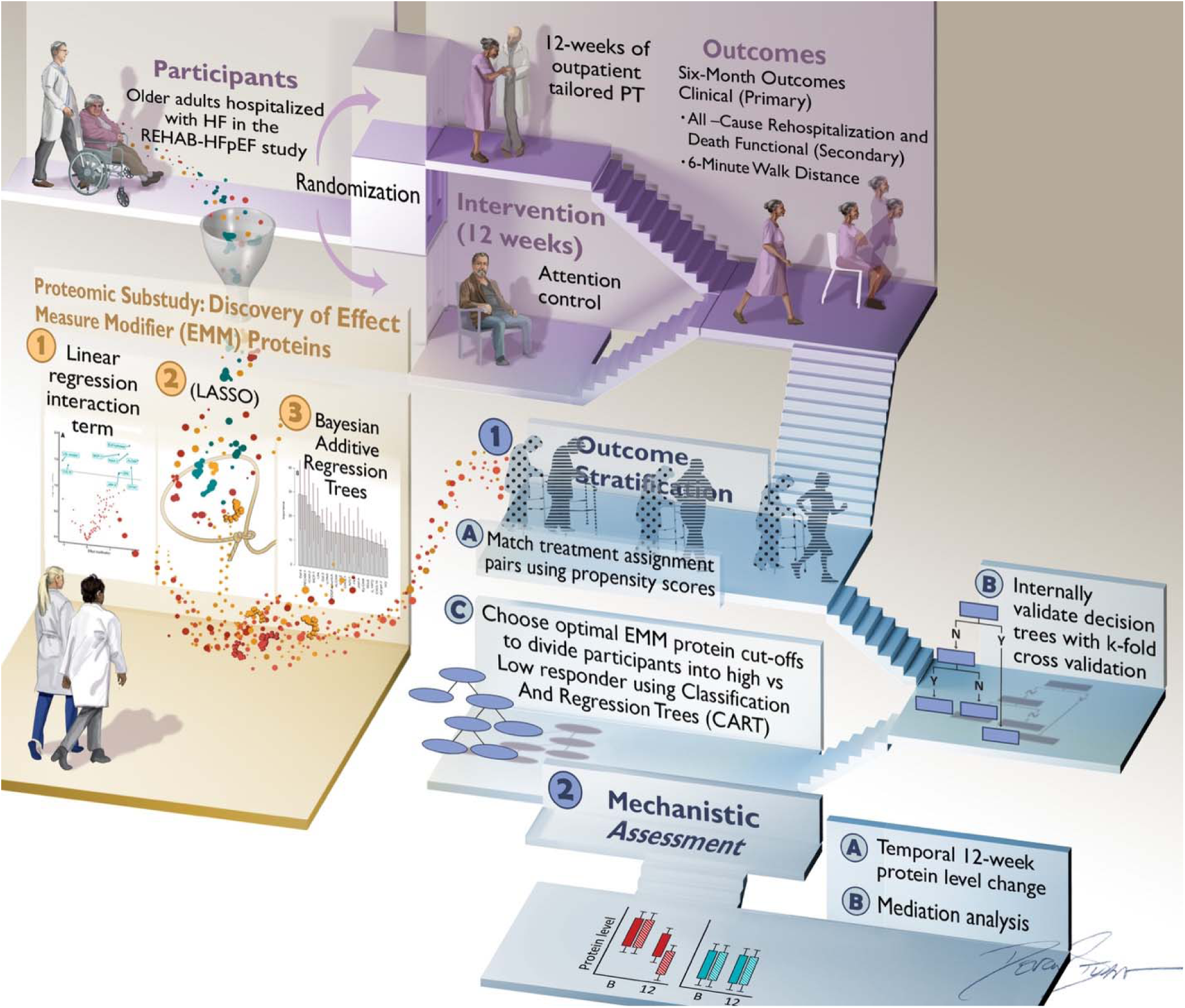
Summary for selection of REHAB-HF participants, randomization to the intervention, study functional outcomes and integration of proteomics with methods for selecting effect measure modifier proteins and decision trees using machine learning methods followed by a causal inference assessment evaluating 12-week change in protein expression levels and their evaluation as mediators to account for change in the functional outcomes. Abbreviations: 6MWD, six-minute walk distance; CART, classification and regression trees; EMM, effect measure modifier; LASSO, Least Absolute Shrinkage and Selection Operator; REHAB-HF, Rehabilitation Therapy in Older Acute Heart Failure Patients.

## Notes

### Competing Interest Statement

The authors have declared no competing interest.

### Clinical Trial

NCT02196038

### Author Declarations

IRB exemption from the University of Maryland. Please see below. Not Human Subjects Research (NHSR) Confirmed To: Christopher deFilippi Link: HP-00120913 Description: An IRB Analyst has reviewed the information provided and has determined that the project meets the definition of Not Human Subjects Research (NHSR). IRB oversight is not required and no further actions are required. Submission Title: Circulating protiens and physical rehabilitation in REHAB-HF POC: Christopher deFilippi Please contact the HRPO at 410-706-5037 or if you have any questions.

